# Assessing the effect of Government of India’s pilot scheme on rice fortification and other etiological factors on the prevalence of anaemia among non-pregnant women in Odisha and Uttar Pradesh

**DOI:** 10.1101/2025.04.30.25326719

**Authors:** Prashanth Thankachan, Boindala Sesikeran, Paramjyoti Chattopadhyay, Ayushi Jain, Vijay Avinandan, Shariqua Yunus, Reema Chugh

## Abstract

Anaemia and micronutrient deficiencies remain pressing public health concerns in developing countries. In India, the prevalence of anaemia among women of reproductive age rose from 53.1% to 57% between 2015–16 and 2019–21, as reported by the National Family Health Survey (NFHS-V). In response, the Government of India launched a pilot scheme in 2019–20 to distribute rice fortified with iron, folic acid, and vitamin B12 through the Targeted Public Distribution System (TPDS) in selected districts. This study evaluates the impact of the pilot scheme after about 20 months of implementation in Malkangiri (Odisha), and 30 months in Chandauli (Uttar Pradesh), using data from a cross-sectional household survey conducted between April and August 2023.

Focusing on non-pregnant women aged 15–49 years, the study aims to: (i) assess changes in anaemia prevalence post-intervention; (ii) examine associations between anaemia and socio-demographic, household, and individual factors; and (iii) identify the most vulnerable groups using Classification and Regression Tree (CART) analysis. NFHS-V data serve as the baseline, and the 2023 survey as the endline. Changes in anaemia prevalence were assessed through descriptive comparisons, while correlates were identified using bivariate statistics and multivariable binary logistic regression. CART analysis was applied to explore pathways to vulnerability.

The findings show a reduction in anaemia prevalence among non-pregnant women in both districts— from 48.5% to 40.9% in Chandauli and 71.8% to 65.7% in Malkangiri. Higher odds of anaemia were observed among scheduled tribe women, those with unimproved sanitation, those not practicing water-tight rice cooking, and those not consuming pulses daily. CART analysis highlighted scheduled tribe women with poor sanitation as the most at-risk group. The study concludes that rice fortification, when combined with improved sanitation and behavioural interventions, offers a promising strategy for reducing anaemia among women of reproductive age in India.

## Introduction

Iron Deficiency Anaemia (IDA) remains a significant health concern globally, particularly among women of reproductive age-group (15-49 years). The World Health Organization (WHO) estimates that anaemia affects half-a-billion women in the reproductive age-group, and the largest causes are iron deficiency, thalassemia and sickle cell trait, and malaria (1). Young women are particularly vulnerable to iron deficiency due to menstrual blood loss and other physiological needs such as pregnancy that results in increased daily iron requirements that needs to be met from the diet, and if the diet fails to provide adequate amounts, a state of iron deficiency (ID) develops leading to IDA if uncorrected. IDA can lead to fatigue, impaired cognitive function, and adverse pregnancy outcomes, posing significant health risks to both women and their children (2).

India, with its diverse population and nutritional landscape, has long grappled with the challenge of addressing malnutrition, including deficiencies in key micronutrients such as iron. WHO estimates that the region of South-East Asia is most affected by anaemia, with 244 million women and 83 million children affected (1), and India sharing a high burden of anaemia. When anaemia is highly prevalent in a population, ID is one of the main contributing nutritional factors besides others in the causal pathway of anaemia. The prevalence of anaemia among all women aged 15-49 years in India, which had decreased between 2005-06 (55.3%) and 2015-16 (53.1%), slipped back to 57% in 2019-21 (3,4). According to WHO’s classification, the latest situation in India falls under the severe category of public health significance (5).

Over the years, there’s been a lot of focus on reducing anaemia globally and India in 2018 has launched a nationwide public health program known as the Anaemia Mukt Bharat (6). The AMB program aims to reduce anaemia prevalence in India by three percentage points every year among six beneficiary groups, including pregnant women, adolescent girls, and children under five. The AMB program has six targeted interventions including, prophylactic iron and folic acid supplementation, deworming, behaviour change communication, anaemia testing using digital hemoglobinometers, provision for fortified foods, and addressing non-nutritional causes of anaemia.

Fortification of staples with micronutrients is a widely accepted complementary strategy and the WHO recognises large scale food fortification as a powerful evidence-informed and cost-effective intervention to address the consequences of vitamin and mineral deficiencies, including iodine deficiency disorders, anaemia and iron deficiency, and neural tube defects among others (7). In the Indian context, fortification of rice is of relevance considering about 65% of India’s population consume rice as staple (8). Fortification programmes across the world are designed in such a manner that they try to bridge the gap between the inadequate dietary intake and estimated average requirements (EAR) or recommended dietary allowances (RDA).

Recognising the reach and the possible effectiveness of rice fortification in reducing the prevalence of anaemia, Government of India launched a pilot scheme beginning 2019-2020 to distribute rice fortified with iron, folic acid and vitamin B_12_ under the Targeted Public Distribution System (TPDS) across fifteen States, of which eleven States reported to roll out the pilot scheme. The scheme was gradually scaled up in the food-based safety net schemes in a phased manner to cover the entire country by 2024, ensuring continuity in the consumption of fortified rice and thus the resulting impact (9). The use of TPDS as a platform to deliver the rice fortification intervention is strategic. Rice is a major TPDS cereal, accounting for about 68% of the cereals distributed in 2023-24 (601.37 lakh MT), followed by wheat (30%) and nutri-cereals (2%) (10). Implying these proportions to the TPDS entitlements (i.e., 5kg of cereals per person for households with a Priority Household [PHH] card; 35 kg per household for those with an Antyodaya Anna Yojana [AAY] card), it is assumed that rice would contribute to about 3kg per person of the cereals distributed under PHH and 24kg per household under AAY, as part of the TPDS distribution setup. As per the latest national estimates, the consumption of rice in India is 5.32 kg per capita/ month in rural and 4.28 kg per capita/ month in urban areas (11).

In India, rice used within the safety net programs is mandatorily fortified with three micronutrients, namely Iron, Folic Acid and Vitamin B_12,_ using extrusion technology for supply in food-based safety net schemes especially to target the problem of nutritional anaemia. Fortified rice is prepared by mixing Fortified Rice Kernel (FRK) with rice in the ratio 1:100. Fortified Rice Kernels (FRK) are made by extrusion technology using rice flour, where micronutrients i.e., iron, folic acid, and vitamin B_12_ are added to rice flour in as per standards prescribed by Food Safety and Standards Authority of India (FSSAI) (12).

Considering a median value of standards, and average consumption to be 100g, fortified rice is expected to meet the EAR of around 24% for iron, 9% for folate and 5% for Vitamin B_12_ among women of reproductive age group. With regards to successful operationalisation of the food fortification programmes, India is uniquely placed as it implements the world’s largest food-based subsidy scheme catering to around 800 million people. The supply chain mechanism for procurement and distribution of wheat and rice is already established and operational. Food Corporation of India (FCI) leads the procurement and supply of food grains including fortified rice to the entire country, while a network of 0.5 million Fair Price Shops ensures the last mile delivery of the fortified rice to beneficiaries. The blending of FRK with normal rice to produce fortified rice is carried out at the level of rice mills are empanelled by the government and from there it is disbursed to the godowns for storage.

While fortified rice is of relevance for the entire country considering pan India high prevalence of anaemia, some Indian states such as Uttar Pradesh and Odisha stand out as regions where anaemia disproportionately impacts women and children. Both these states were selected to implement pilot scheme on fortification of rice and its distribution under TPDS as well. Since the pilot was proposed for rolling out in one district per state, majority of the states including UP and Odisha, selected the most vulnerable, rice producing and consuming districts as per the NITI Aayog’s list of aspirational districts. In UP, the pilot scheme was implemented in the district of Chandauli from January 2021 while in Odisha, the pilot was rolled out in the tribal dominated district of Malkangiri since July 2021. It took about 2-3 months in both Chandauli and Malkangiri districts to fully optimise the supply chain for distribution of fortified rice, ensuring non fortified rice is replaced with fortified rice across all Fair Price shops in the districts and achieve a coverage of 100% in terms of beneficiary reach.

Post the roll out of the pilot scheme on rice fortification under TPDS, an assessment was undertaken by World Food Programme in Chandauli and Malkangiri, to study the impact of the pilot scheme on the prevalence of anaemia among four demographic groups including children (6-59 months), adolescents (15-19 years), women (20-49 years), and men (20-49 years) in the TPDS availing households. Since the rice fortification scheme was rolled out closer to the fifth round of National Family Health Survey-5 in both the districts, NFHS-5 findings were considered as baseline and this assessment as endline for comparison. The endline assessment was conducted to assess the effect of consumption of fortified rice in the two districts of Malkangiri and Chandauli after 20 months and 30 months respectively of fortified rice distribution of the implementation of the rice fortification scheme. However, as far as the scope of this paper is concerned, we focus solely on reproductive age women aged 15-49 years who were not pregnant at the time of survey.

The paper attempts to fulfil three objectives. First, it provides an overview of rice fortification programme in terms of its scope, coverage, and implementation across the two districts and the households covered in the survey. Second, it assesses the change in the prevalence of anaemia among non-pregnant women (15-49 years) after the introduction of the rice fortification pilot scheme in the district comparing with NFHS-5 data. Third, it examines the intersectionality between anaemia prevalence and various socio-economic, demographic, and individual factors in non-pregnant women of reproductive age (15-49 years). It also identifies the most vulnerable groups using Classification Analysis & Regression Trees (CART).

## Materials and Methods

### Data source and Sampling

The study uses cross-sectional data from 968 households in Chandauli and 1056 households in Malkangiri across 44 randomly drawn Primary Sampling Units (PSUs) split between rural and urban areas, covered during the primary data collection as part of endline assessment. The inclusion criterion was households receiving fortified rice through the TPDS or state food security programme. The endline assessment used a two-stage stratified sampling approach to select households, same as that of the National Family Health Survey-5 sampling methodology (4). Census 2011 data was used as the sampling frame for the selection of PSUs. Villages in rural areas and Census Enumeration Blocks (CEBs) in urban areas were considered as PSUs. In the first stage of sampling, a fixed number of PSUs were selected from both rural and urban strata in each district using the Probability Proportional to Size (PPS) stratified sampling method, with 48 PSUs selected in Malkangiri (44 rural and 4 urban) and 44 PSUs in Chandauli (38 rural and 6 urban). Before selecting the PSUs, each rural stratum was divided into six approximately equal substrata, based on the proportion of households in each PSU and the percentage of the population from Scheduled Castes (SC) and Scheduled Tribes (ST). Within each rural sampling stratum, PSUs were sorted by the literacy rate of women aged 6 years and above. The rural PSUs were then sampled using the PPS method. For PSUs with more than 300 households, they were subdivided into segments of roughly 100-150 households, and two segments were randomly selected for the survey through systematic sampling, with selection probability based on segment size. In the second stage, 22 households per PSU were randomly selected using systematic sampling from a newly created list of eligible households (derived from house listing) in the chosen PSUs.

The TPDS/Anganwadi beneficiary lists served as the sampling frame for selecting the 22 households. For the urban PSUs, selection was based on the 2011 census wards. All urban wards were sorted by the percentage of SC/ST population, and the 4 PSUs in Malkangiri district and 6 PSUs in Chandauli district were chosen using the PPS method. After selecting the wards, the same household listing procedure was applied to select households in urban areas.

### Target Group

The endline assessment covered a total of 1265 women in Chandauli and 1095 women in Malkangiri in the reproductive age group (15-49 years) whose blood haemoglobin was measured. The inclusion criteria required selecting at least one woman aged 20-49 from each household, along with one or more adolescent girls aged 15-19, either from the same household or from a different one, based on their availability in the selected household. Oral assent was obtained from minor participants (<18 years of age) and written informed consent was taken from the adults (>18 years of age) before conducting interviews and collecting blood specimens.

### Tool Used

The women were interviewed using a structured tool that gauged the household’s access, utilization, and acceptability of the fortified rice received under TPDS in addition to collecting information on some socio-demographic indicators. The survey also collected individual-level data on various factors, such as anaemia prevalence, regular access to and acceptability and consumption of fortified rice, Iron and Folic Acid (IFA), and iron-rich foods. Dietary assessments were also undertaken using NFHS-5 dietary questionnaire based on 24 hours recall period. The questionnaire was administered in their local language in both the districts.

### Haemoglobin Testing

The study employed the capillary blood testing method for haemoglobin estimation which was the same method used in NFHS-5. Blood samples were drawn from a drop of blood taken from a finger prick and collected in a microcuvette. Haemoglobin analysis was conducted on-site with a battery-operated portable analyser. Two types of analysers were used: HemoCue in Chandauli and AccuSure in Malkangiri. Comparison study for both analysers was conducted in accredited laboratories prior to the study implementation to ensure comparability.

### Ethics

The study was conducted in full compliance with ethical guidelines and practices, with a strong emphasis on participant confidentiality and well-being throughout fieldwork and data collection. Participants were informed that their involvement was completely voluntary, and their personal information would remain confidential. Written informed consent was obtained from all participants. For minor participants, and informed consent was secured from their parents prior to interviews and blood specimen collection. Study materials, including information sheets and consent forms, were provided in both Hindi and English.

Participants’ identities were carefully protected, and all data was anonymized to ensure privacy. The research team also demonstrated respect for local customs and cultural norms, adopting culturally appropriate conduct to prevent any negative emotional impact on participants. Ethical approval for this study was granted by the Institutional Review Board (IRB) of TRIOs, a body registered with the Office for Human Research Protections (OHRP) under the U.S. Department of Health and Human Services (HHS).

### Survey Period

Data collection in Malkangiri district started on 1 April 2023 and ended on 15 May 2023, while in Chandauli district, the data collection started on 1 August 2023 and ended on 31 August 2023. This occurred after about 20 months and 30 months of implementing the rice fortification scheme in the respective districts.

### Comparison Data

To compare the change in anaemia prevalence in the endline, we used the haemoglobin levels of non-pregnant women (aged 15-49) recorded in the NFHS-5 (2019–21) from both districts as the baseline.

### Analytical Sample

The cross-sectional data of non-pregnant women aged 15-49 years from the Chandauli and Malkangiri districts was pooled after screening for common variables with available data for chi-square and regression analysis. Respondents with sickle cell disease or thalassemia were excluded from the study. A complete case analysis was performed, meaning that missing observations were meticulously examined and excluded. The unit of analysis is household and to ensure data integrity, only one randomly selected respondent per household was included to avoid multiple counts of household-level variables in the analysis. This approach avoids inflated or biased estimates in the logistic regression model. Using this approach, a final analytical sample consisting of information on 1865 respondents was obtained. Additionally, sensitivity analysis was performed to validate the main findings. Respondents with same household IDs who were excluded in the final analytical sample were replaced with their counterparts. The same statistical tests were then performed, and the regression model were compared to identify any observable differences.

The detailed steps carried out to achieve the final analytical sample and for sensitivity analysis is presented in Fig 1.

**Fig 1:**
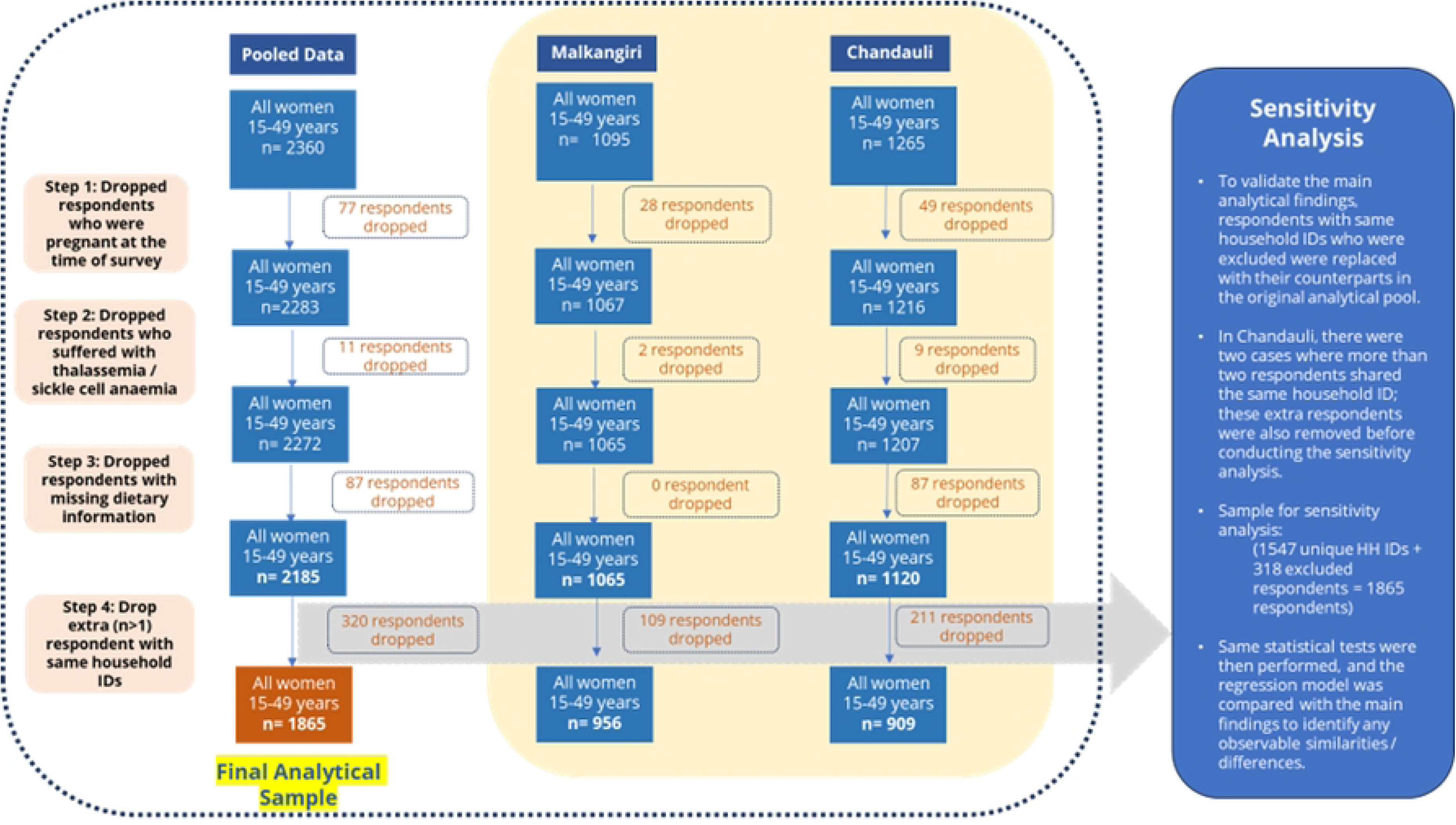
Schematic showing the steps undertaken to obtain the final analytical sample and sensitivity analysis.

## Study framework

The analysis for this study is based on a conceptual framework adapted from Lancet global framework on malnutrition (13). The framework depicted in Fig 2 highlights various anaemia-related factors that could influence the prevalence of anaemia in the selected districts. Three key categories of variables were chosen for the regression analysis: household, individual and diet & supplementation. These are detailed in the following section.

**Fig 2:**
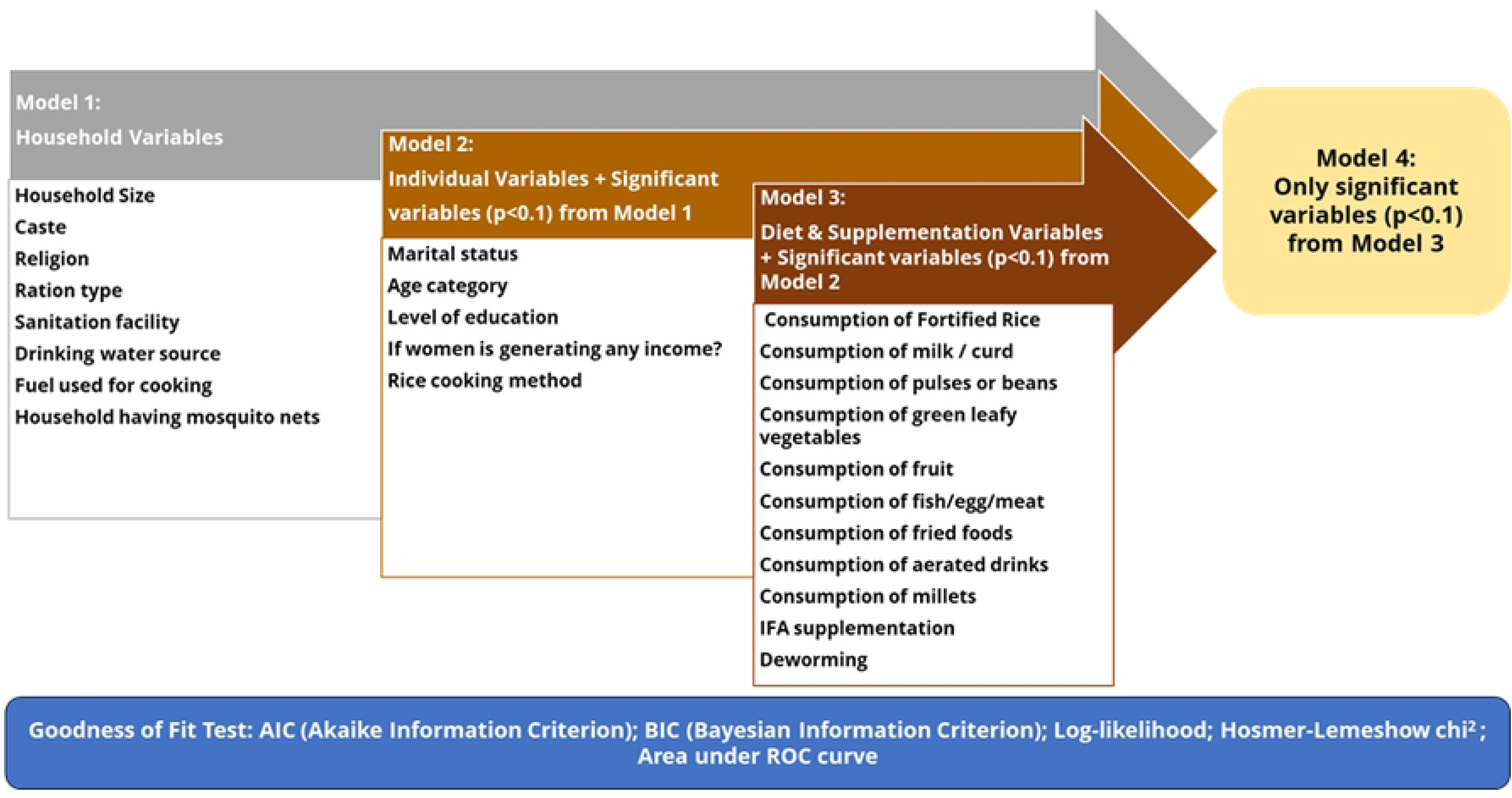
Conceptual framework for adjusted regression analysis

### Dependent Variable

The dependent variable in this study was whether non-pregnant women in the reproductive age-group were anaemic or not. It was coded as a dichotomous variable, where ‘1’ represents women who are anaemic (including mild, moderate, or severe anaemia), and ‘0’ represents women who are not anaemic. For non-pregnant women, anaemia is classified as ‘mild’ if the haemoglobin level ranges from 11.0 g/dl to 11.9 g/dl. It is categorized as ‘moderate’ if the haemoglobin level is between 8.0 g/dl and 10.9 g/dl. Anaemia is considered ‘severe’ if the haemoglobin level is below 8.0 g/dl. Non-pregnant woman is ‘not anaemic’ if haemoglobin level is equal or greater than 12.0 g/dl.

### Independent Variable

Table 1 provides a detailed description of independent variables used in the study, for which the data was collected from the respondents during the household survey.

**Table 1:** Description of independent variables.

| S. No. | Category | Name of Variable | Description | Recoded as |
| --- | --- | --- | --- | --- |
| 1 | Demographic | District | Name of district | 0. Chandauli<br>1. Malkangiri |
| 2 | Demographic | Household Size | Total number of members in family | 0. 1-4 members<br>1. 5-6 members<br>2. 7+ members |
| 3 | Demographic | Caste | Social category of the household | 0. OBC/General<br>1. SC<br>2. ST |
| 4 | Demographic | Religion | Religion of the household | 0. Hindu<br>1. Other |
| 5 | Demographic | Ration card type | Type of ration card owned by the household | 0. PHH<br>1. AAY |
| 6 | Household | Drinking water facility | Main source of drinking water for members of your household? | 0. improved source (piped drinking water, public taps, standpipes, tubewells, boreholes, protected dug wells and springs, rainwater, tanker truck, cart with small tank, bottled water, community reverse osmosis plants)<br>1. not improved source (any other source) |
| 7 | Household | Sanitation facility | Kind of toilet facility do members of household usually use | 0. improved facility (flush/ pour toilets to piped sewer system, septic tanks, pit latrines, or unknown destination; ventilated improved pit/ biogas latrines; pit latrines with slabs; twin-pit/ composting toilets)<br>1. unimproved facility (any other source) |
| 8 | Household | Type of cooking fuel | Type of fuel does your household mainly use for cooking | 0. clean fuel (electricity, LPG/ natural gas, biogas)<br>1. unclean fuel (any other source) |
| 9 | Household | Households have mosquito net | Household having any mosquito nets | 0. yes<br>1. no |
| 10 | Individual | If woman is generating any income? | If woman is engaged in any occupation | 0. yes<br>1. no |
| 11 | Individual | Marital status | Marital status of woman | 0. currently married<br>1. not married (never/divorced/widow) |
| 12 | Individual | Age category | Woman's age in years | 0. 15-19 years<br>1. 20-24 years<br>2. 25-34 years<br>3. 35-49 years |
| 13 | Individual | Education level | Woman's education level | 0. no schooling<br>1. primary<br>2. secondary<br>3. graduate and above |
| 14 | Individual | Rice cooking method | Rice cooking method | 0. watertight method<br>1. openvessel_dontknow |
| 15 | Diet & Supplementation | Consumption of Fortified Rice | Frequency of consuming fortified rice in a week | 0. daily<br>1. notdaily |
| 16 | Diet & Supplementation | Consumption of milk / curd | Frequency of consuming Milk or curd in a week | 0. daily<br>1. notdaily |
| 17 | Diet & Supplementation | Consumption of pulses or beans | Frequency of consuming Pulses or beans in a week | 0. daily<br>1. notdaily |
| 18 | Diet & Supplementation | Consumption of green leafy vegetables | Frequency of consuming Dark green leafy vegetables in a week | 0. daily<br>1. notdaily |
| 19 | Diet & Supplementation | Consumption of fruit | Frequency of consuming fruits in a week | 0. daily_weekly<br>1. occasionally_never |
| 20 | Diet & Supplementation | Consumption of fish/egg/meat | Frequency of consuming either Fish or Eggs or Chicken or meat in a week | 0. daily_weekly<br>1. occasionally_never |
| 21 | Diet & Supplementation | Consumption of fried foods | Frequency of consuming fried foods in a week | 0. daily_weekly<br>1. occasionally_never |
| 22 | Diet & Supplementation | Consumption of aerated drinks | Frequency of consuming aerated drinks in a week | 0. daily_weekly<br>1. occasionally_never |
| 23 | Diet & Supplementation | Consumption of millets | Frequency of consuming millets in a week | 0. daily_weekly<br>1. occasionally_never |
| 24 | Diet & Supplementation | Received IFA supplement in last 7 days? | Was IFA tablets or syrup been given seven days prior to the survey? | 0. yes<br>1. no |
| 25 | Diet & Supplementation | Received deworming tablet in last 6 months? | Was any drug for intestinal worms been given in six months prior to the survey? | 0. yes<br>1. no |

### Statistical analysis

The paper first captures in detail the roll-out and implementation of fortified rice intervention in Chandauli and Malkangiri districts, followed by description of households covered in both the districts. Further, trends in anaemia prevalence and severity among non-pregnant women were examined before and after the rollout of the fortified rice scheme in both districts, using NFHS-5 (2019–21) as the baseline and the current study as the endline. Descriptive statistics along with Pearson Chi-square test was employed to study the mean prevalence and to assess the statistical significance of the relationship between the outcome variable and each independent variable. Furthermore, unadjusted regression was done to examine the influence of individual variables on the odds of anaemia without accounting for any confounding variables. Then, adjusted binary logistic regression analyses were conducted following a conceptual framework presented in Figure 2. Relevant variables from each level were selected in a sequential manner to identify significant predictor variables, treating anaemia as outcome variable. The analyses were conducted in four steps. First, the variables from household category were added in the model (adjusted model 1), then significant variables (p<0.1) from this category were added along with individual category variables (adjusted model 2). Significant predictors from model 2 were then added along with variables under diet & supplementation category (adjusted model 3). Finally, only the significant predictors from model 3 were adjusted together in a Model 4. This stepwise introduction of predictor variables ensures the consistency of their performance and helps identify those with the most substantial influence on the outcome variable i.e. anaemia. Models were assessed using goodness-of-fit tests (AIC, BIC, Log-likelihood, Hosmer-Lemeshow chi-square, area under ROC curve). Higher log likelihood, and low AIC and BIC values indicated a better fit. The Hosmer-Lemeshow test was used to assess how well our model’s predictions matched the actual outcomes, and if the result were not significant (p>0.05), it suggested a good fit. For area under ROC curve, value of 0.6 suggested moderate discrimination, while 0.7 suggested acceptable discrimination.

Further, CART was conducted to identify the most vulnerable groups. The CART tree is produced through an algorithm that identifies the groups that have different prevalence rates that are statistically significant. In doing so, it identifies the furthest behind and the furthest ahead groups. Moving from left to right, the population is split into smaller groups if the splitting criterion allows, or until the sample size becomes too small. Only circumstances that show significant variance are picked up by the algorithm and subsequently feature in the trees. Predictors from the adjusted model 4 of the regression analysis were used as ‘circumstances’, while ‘prevalence of anaemia’ was considered as ‘outcome variable’. Other parameters considered included complexity parameter = 1; minimum splitting criteria = 0.09; minimum sample size=49; method = ANOVA.

## Results

### Overview of rice fortification programme in terms of its scope, coverage, and implementation in surveyed districts

In Uttar Pradesh, the pilot scheme was implemented in the district of Chandauli from January 2021 while in Odisha, the pilot was rolled out in the tribal dominated district of Malkangiri since July 2021, each district taking almost 2-3 months to fully stabilise and reach saturation. Under TPDS, food grains are provided to two categories of beneficiaries, the priority households (PHH) and Antyodaya Anna Yojana (AAY) families. All priority households are entitled to receive food grains at rate of 5kg per beneficiary per month while Antyodaya families (AAY) receive 35kg of grains per household per month. The districts of Chandauli and Malkangiri are culturally very different and have differential preference for food commodities provided through the TPDS. In Malkangiri, rice is a staple food and is the only food grain supplied through TPDS, while in Chandauli, beneficiaries are provided both wheat and rice. During the roll out of the pilot scheme, around 0.45 million beneficiaries of TPDS in Malkangiri received 100% fortified rice while in case of Chandauli, fortified rice was introduced in a staggered manner over the due course of the roll out. In Chandauli, at the start of the programme, from January 2021 to May 2022, each PHH beneficiary was provided 3 Kg wheat and 2 Kg fortified rice. However, from June 2022, this has changed to 3 kg fortified rice and 2 kg wheat. For AAY beneficiaries, the ratio was maintained at 21 kg fortified rice and 14 kg wheat. The total number of beneficiaries reached with fortified rice in Chandauli stood at around 1.4 million. Other than TPDS, fortified rice was also distributed in other food safety net schemes of PM POSHAN and Anganwadi Service Schemes in both the states. In Chandauli, for PM-POSHAN, the distribution of fortified rice started in February 2021 and for Anganwadi Service Schemes in March 2021, while in Malkangiri, the distribution in both the schemes started in October 2021. In terms of procurement, Odisha (Malkangiri) follows a decentralised procurement model and uses the fortified rice produced with in the state itself, while in Uttar Pradesh (Chandauli), the state follows a centralised model where FCI procures all the fortified rice produced within the state and then depending on its requirement, state procures the desired quantity from FCI. Prior to the roll out of the scheme in 2021, both the states invested hugely in creating desirable infrastructure within the selected districts to ensure continuous supply of adequate and quality fortified rice. In Malkangiri, 28 blending equipment were installed and the total blending capacity for the district is 724.1 MT per 8 hours against the average monthly requirement of around 2800 MT while in Chandauli, 48 blending equipment were in operation with a total blending capacity for the district is 2201 MT per 8 hours against the average monthly requirement of around 5041 MT. In terms of ensuring quality production and supply of fortified rice, both the districts followed guidelines as stipulated by the Department of Food and Public Distribution, wherein it was mandated for FRK manufacturers to get each batch of FRK tested for micronutrient and microbiological parameters and share the copy of test result (Certificate of Analysis) with the rice millers and the state government. This was followed by testing of the sample of fortified rice for blending efficiency at the time of receipt of fortified rice at the storage godown. Ten percent of the fortified rice stored in the godown were also tested for micronutrient content at an NABL accredited lab by procurement agency.

### Characteristic of households covered in the survey in both districts

Majority (72.4%) of the households in Malkangiri belonged to scheduled tribes while in Chandauli, only 4.5% households belonged to scheduled tribes and majority (58.1%) belonged to other backwards classes and other. Nearly 37.4% households belonged to scheduled caste in Chandauli in comparison to only 15.2% in Malkangiri. Unlike caste, majority of the households in both Malkangiri (98.9%) and Chandauli (92.3%) districts belonged to Hindu religion. A comparison of the family size between the two districts shows that around 70% households in Chandauli had a family size of more than 4 members, whereas in Malkangiri, nearly 50% of the households interviewed had a family size of more than 4 members. In Chandauli, nearly 85% households interviewed owned a Priority Household (PHH) ration card and 15% had AAY card, while in Malkangiri, 91.4% households were PHH card holders and only 8.7% were AAY card holders. In terms of access to basic amenities such as drinking water, clean fuel, sanitation facility, access and use of mosquito nets which indirectly affects anaemia prevalences across age groups, 97.6% households in Chandauli had access to improved drinking-water source and 79.6% households had access to improved sanitation facility while in Malkangiri, 92.6% households had access to improved drinking-water source but only 26.3% households had access to improved sanitation facility. Access to clean fuel appeared as a major barrier with more than 50% households in Chandauli and around 80% households in Malkangiri not having access to clean fuel for cooking. Sixty five percent households in Chandauli and 85% in Malkangiri had access to mosquito nets and majority of them were using the mosquito nets (Table 2).

**Table 2:** Characteristics of households covered in the endline assessment in Chandauli and Malkangiri district.

| Variable | Category | Chandauli, N (%) | Malkangiri, N (%) |
| --- | --- | --- | --- |
| Household's Caste | Scheduled Caste | 362 (37.4) | 160 (15.2) |
|  | Scheduled Tribe | 44 (4.5) | 763 (72.4) |
|  | Others (OBC, General) | 562 (58.1) | 131 (12.4) |
| Household's Religion | Hindu | 893 (92.3) | 1042 (98.9) |
|  | Others (Muslim, Christian, Others) | 75 (7.7) | 12 (1.1) |
| Number of household members | ≤4 | 283 (29.2) | 513 (48.7) |
|  | 5 to 6 | 453 (46.8) | 435 (41.3) |
|  | >6 | 232 (24.0) | 106 (10.1) |
| Household's living conditions | Population living in households with an improved drinking-water source | 845 (97.6) | 976 (92.6) |
|  | Population living in households that use an improved sanitation facility | 771 (79.6) | 277 (26.3) |
|  | Households using clean fuel for cooking | 437 (45.1) | 195 (18.6) |
| Type of Ration Card | PHH | 821 (84.8) | 963 (91.4) |
|  | AAY | 147 (15.2) | 91 (8.7) |
| Mosquito Nets | Households having mosquito nets | 637 (65.8) | 898 (85.2) |
|  | Households using mosquito nets | 531 (54.9) | 557 (52.8) |

### Shift in prevalence of anaemia among non-pregnant women between baseline and endline

Compared to the baseline (NFHS-5), the endline prevalence of anaemia significantly declined in both the districts (p<0.05), however the net change in the median haemoglobin levels were found to be negligible. The prevalence of ‘any anaemia’ declined by 7.6 percentage points in Chandauli (from 48.5% to 40.9%) while in Malkangiri, a 4.6 percentage points (from 71.8% to 67.2%) decline was observed. In terms of severity, both Chandauli and Malkangiri showed statistically significant decline in prevalence of ‘severe’ and ‘moderate’ anaemia, with maximum decline being observed in ‘moderate’ anaemia. While an increase in ‘mild’ anaemia prevalence was observed among non-pregnant women in both Malkangiri and Chandauli district in the endline (Table 3).

**Table 3:** Shift in prevalence of anaemia by severity among non-pregnant women (15-49 years) in Chandauli and Malkangiri district between NFHS-5 and endline

| Description | Non-pregnant women (15-49 years) |  |  |  |  |  |
| --- | --- | --- | --- | --- | --- | --- |
|  | Chandauli |  |  | Malkangiri |  |  |
|  | NFHS-5<br>(2019-21) | Endline<br>(2023) | Statistical<br>difference<br>(p-value) | NFHS-5<br>(2019-21) | Endline<br>(2023) | Statistical<br>difference<br>(p-value) |
| Weighted Sample (n) | 1071 | 1210 | na | 358 | 1053 | na |
| Median Hb levels, in g/dl | 12 | 12.1 | 0.019 | 11.2 | 11.3 | 0.216 |
| Any anaemia (<12.0 g/dl), in % | 48.5 [45.2 - 51.7] | 40.9 [37.6 - 44.4] | 0.002 | 71.8 [68.9 - 74.5] | 67.2 [63.7 - 70.5] | 0.04 |
| Severe Anaemia (<8 g/dl) in % | 1.5 [.9 - 2.4] | 0.2 [0.1 - 0.7] | 0.002 | 2.2 [1.5 - 3.4] | 0.5 [0.2 - 1.2] | 0.003 |
| Moderate Anaemia (8-10.9 g/dl), in % | 22.8 [20.1 - 25.7] | 6.9 [5.5 - 8.7] | 0.000 | 41.5 [38.4 - 44.5] | 21.5 [18.9 - 24.4] | 0.000 |
| Mild Anaemia (11-11.9 g/dl), in % | 24.2 [21.5 - 27.1] | 33.8 [30.5 - 37.1] | 0.000 | 28.1 [25.4 - 31.0] | 45.1 [41.6 - 48.6] | 0.000 |

### Descriptive statistics and Chi-square tests

The Chi-square test results for pooled data using the final analytical sample (n=1865) (Table 4) revealed significant differences in anaemia prevalence among non-pregnant women of reproductive age between the two districts, with 69.7% of women in Malkangiri being anaemic compared to 43.7% in Chandauli. Among household variables, caste played a key role; anaemia was highest among scheduled tribes (68.9%), followed by scheduled castes (51.9%) and OBC/general categories (47%). Ration card type also mattered, with women from Priority Households (PHH) having higher anaemia prevalence (57.9%) than those from Antyodaya Anna Yojana (AAY) households (50.2%). Women using unimproved drinking water sources had a significantly higher prevalence (68.8%) compared to those using improved sources (56.5%). Similarly, access to sanitation affected anaemia, with women lacking improved sanitation having higher prevalence (66.5%) compared to those with improved facilities (47.4%). The use of unclean cooking fuel was linked to higher anaemia prevalence (61.2%) compared to clean fuel (48.1%). In terms of individual factors, women not involved in any income-generating activity had a lower anaemia prevalence (54.4%) compared to those who did (67.7%). Education also played a major role-women with no schooling had the highest prevalence (63.8%), while those with secondary education (51.3%) and graduates (41.6%) fared better. Cooking methods impacted anaemia, with the watertight cooking method linked to lower prevalence (43.4%) compared to open vessel methods (61.6%). Dietary patterns also show a significant association-daily milk or curd consumption was associated with lower anaemia (46.6% vs. 58.3%); the same held true for daily pulses or beans consumption (52.3% vs. 64.1%) and frequent fruit consumption (50.9% vs. 59.3%).

**Table 4:**
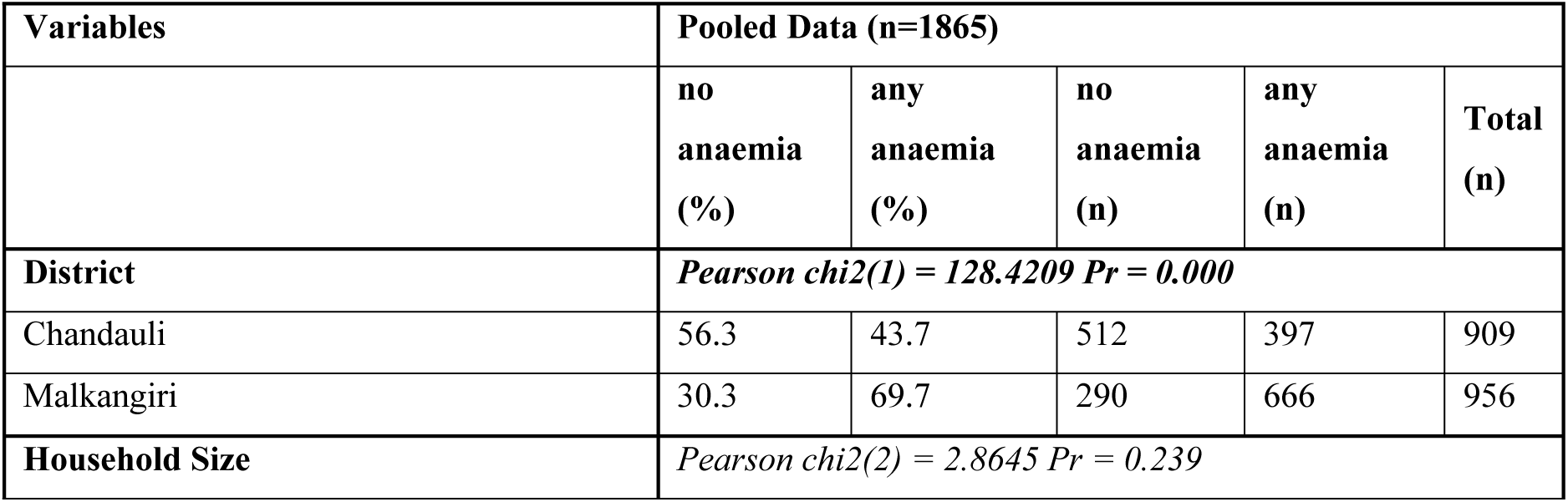
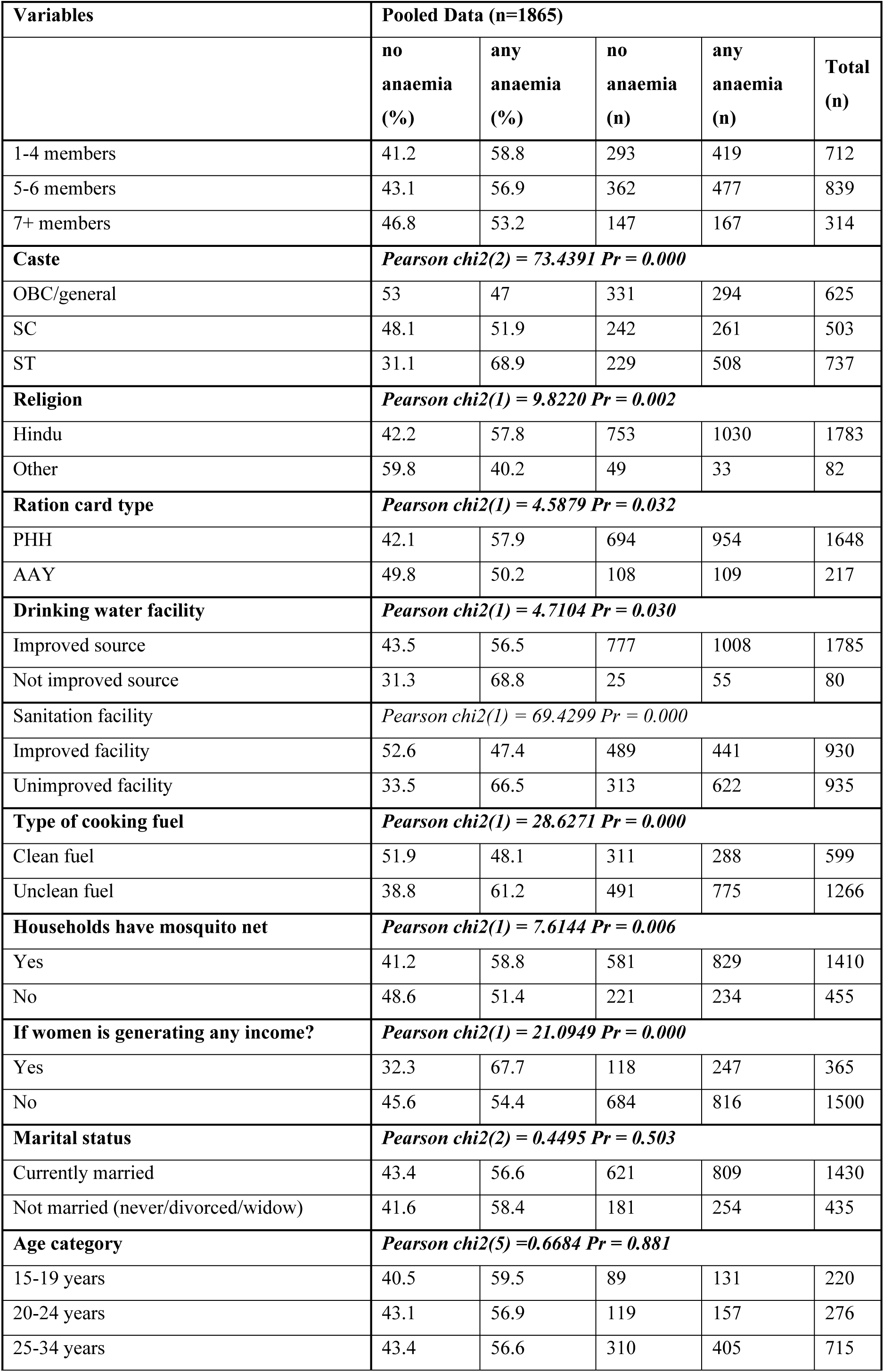
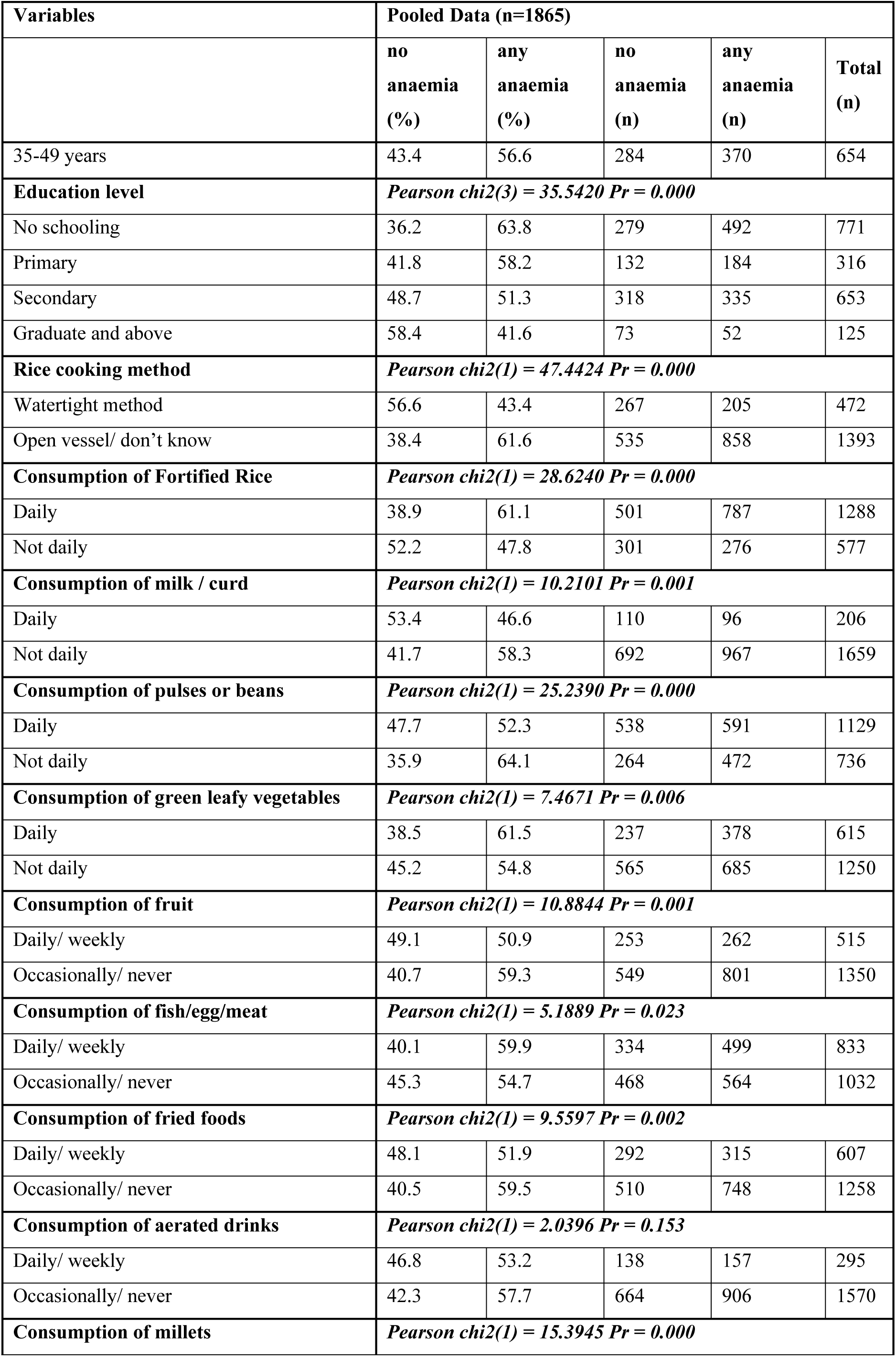
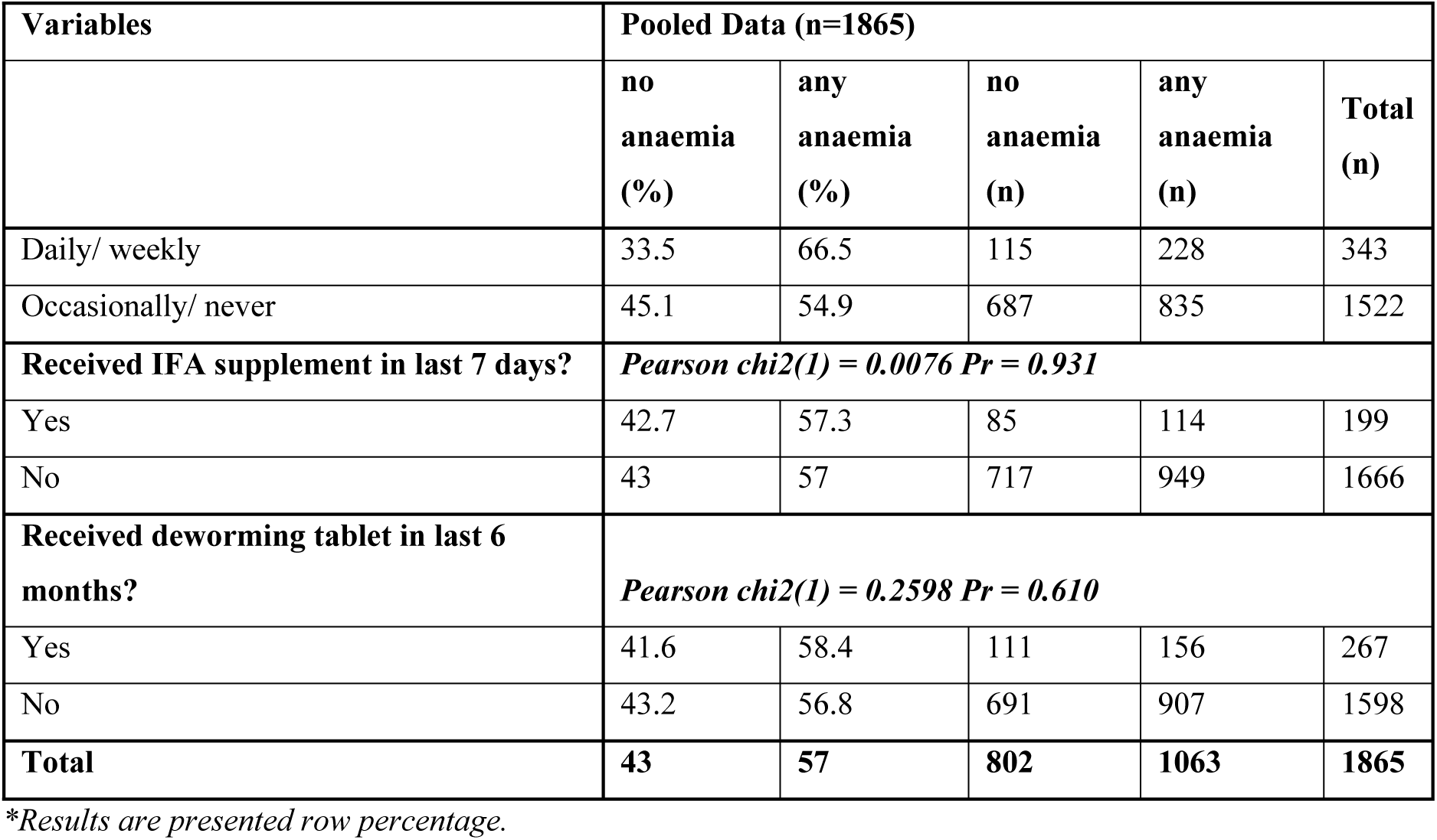
Descriptive Statistics and Chi-square results.

Despite the improvement in anaemia status among non-pregnant women, non-pregnant women who reported to consume fortified rice daily had higher anaemia prevalence (61.1%) due to poor socio-economic background, poor living conditions and poor dietary intake as observed in further analysis presented in Supplementary Table 2 and detailed in the discussion section later. Frequent millet consumption also showed an association with higher anaemia (66.5% vs. 54.9%). The explanation for these counter-intuitive findings is further elaborated in the discussion section.

### Adjusted Regression Analysis

As observed in Table 5, the goodness-of-fit test indicates improvement of the model as we eventually proceed to adjusted model 4 using the pooled data, confirming the better model prediction with inclusion of predictors in sequential manner. The adjusted model 4 highlight several variables significantly associated with anaemia prevalence among non-pregnant women of reproductive age (15-49 years). Women from scheduled tribes had 58 percent higher odds of being anaemic compared to women from OBC/General categories (AOR: 1.58, 95 percent CI: 1.22-2.06). Similarly, access to basic services plays a critical role-women from households without improved sanitation facilities had 59 percent higher odds of anaemia (AOR: 1.59, 95 percent CI: 1.28-1.98) compared to those with improved sanitation. Women from Antyodaya Anna Yojana (AAY) ration card households had 29 percent lower odds of anaemia compared to those from Priority Household (PHH) families (AOR: 0.71, 95 percent CI: 0.53-0.95). The method of cooking also showed a borderline association, with women using the open vessel method having 27 percent higher odds of anaemia compared to those using the watertight method (AOR: 1.27, 95 percent CI: 0.99-1.63). Dietary habits were also significant factors in determining anaemia prevalence. On the other hand, women who did not consume pulses or beans daily had 26 percent higher odds of anaemia (AOR: 1.26, 95 percent CI: 1.02-1.55) compared to those who consumed them daily. Women’s age had a borderline effect, as women aged 25-34 years had 24 percent lower odds of anaemia compared to the adolescents (AOR: 0.76, 95 percent CI: 0.55-1.05). It was also found that women who did not consume fortified rice daily had 22 percent lower odds of being anaemic compared to daily consumers (AOR: 0.78, 95 percent CI: 0.63-0.97), which presents a counter-intuitive result and has been discussed later.

**Table 5:**
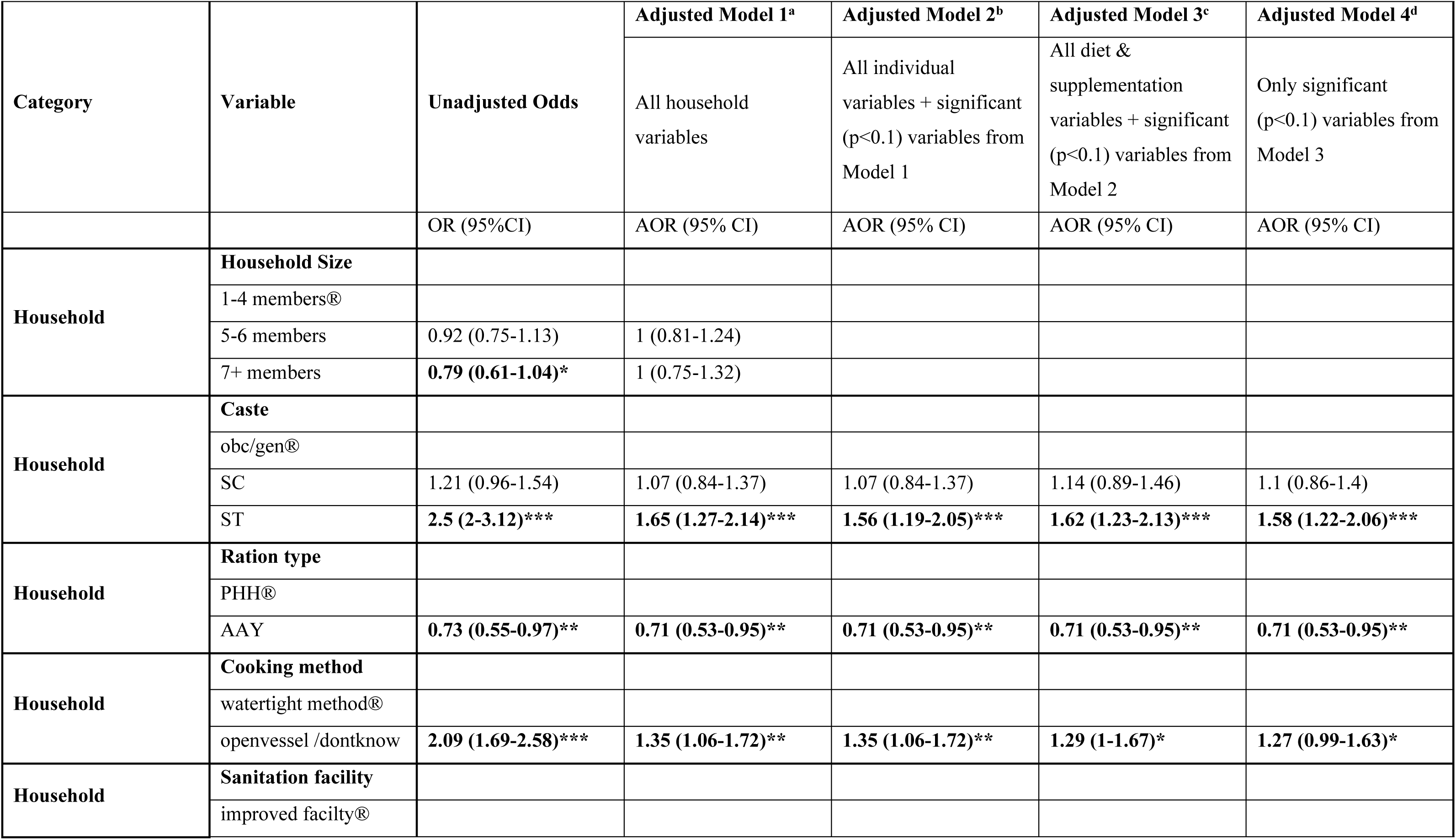
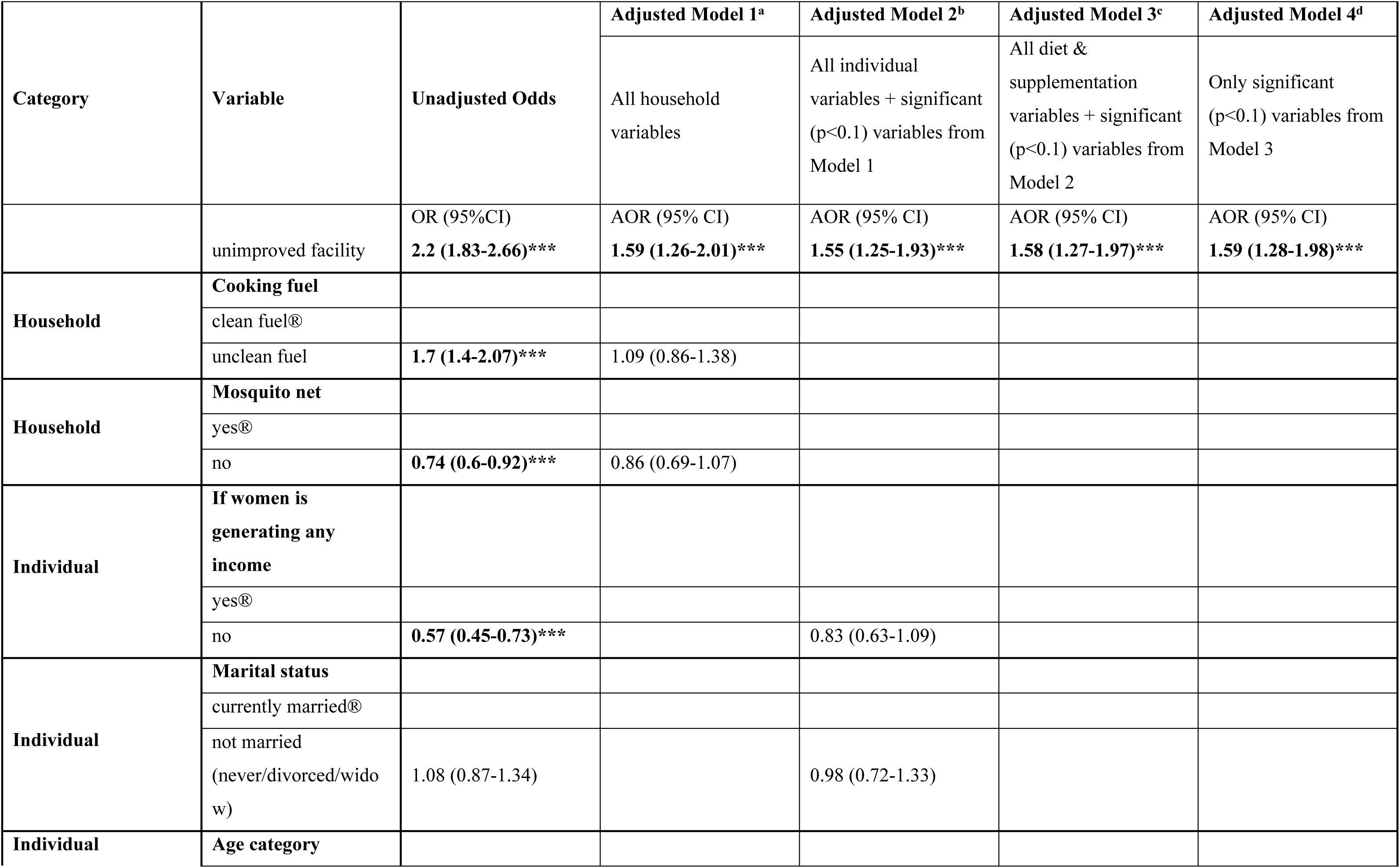
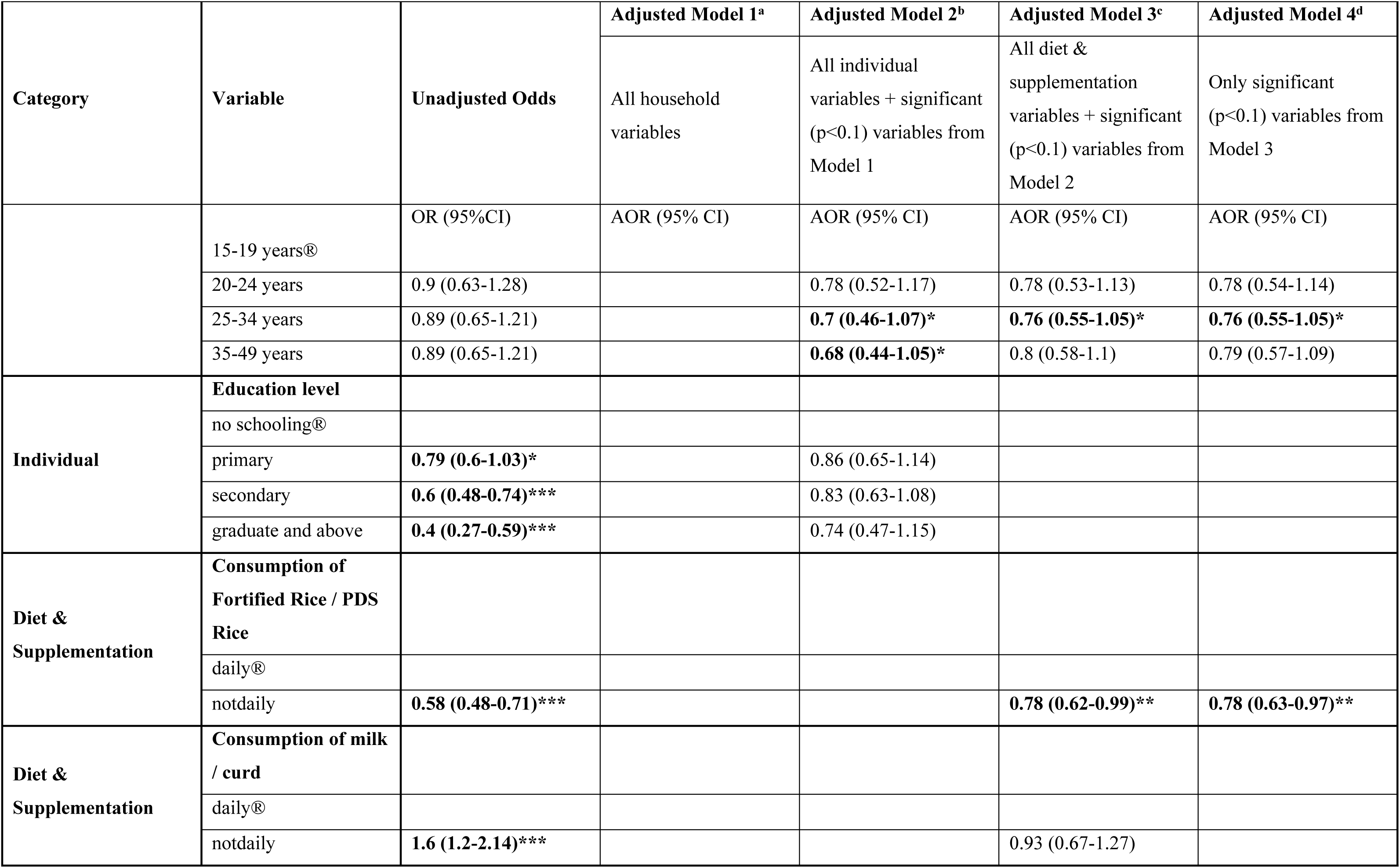
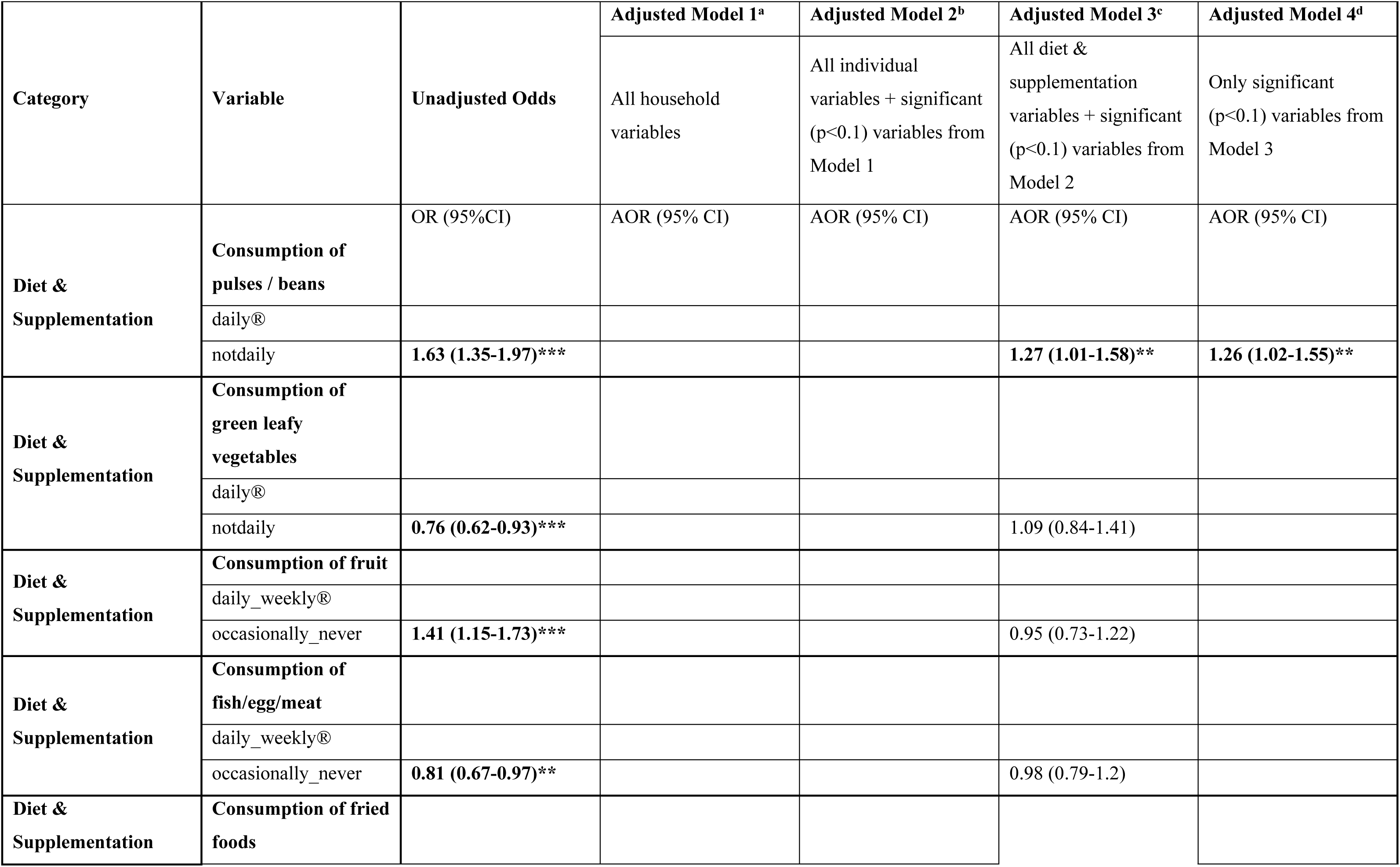
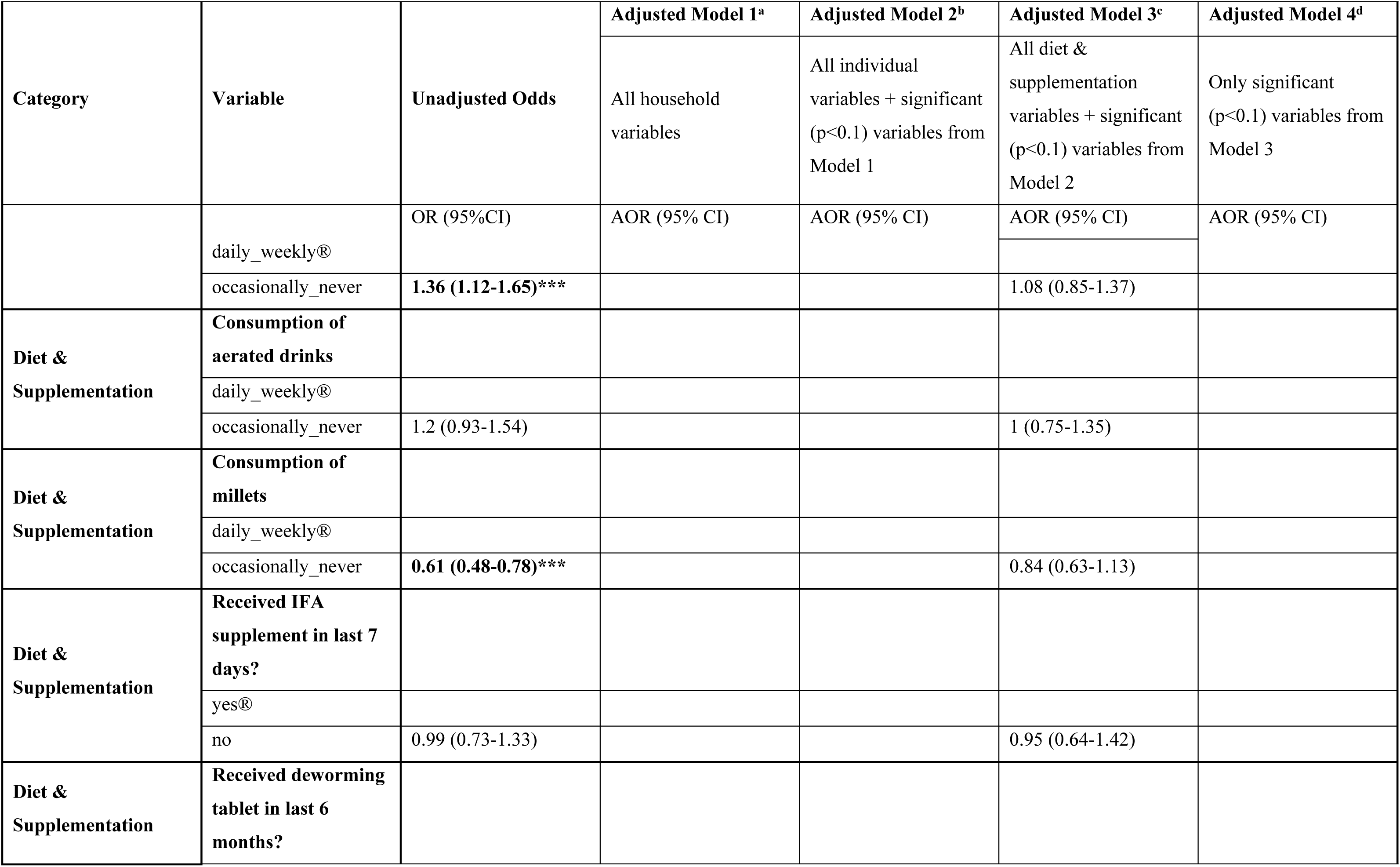
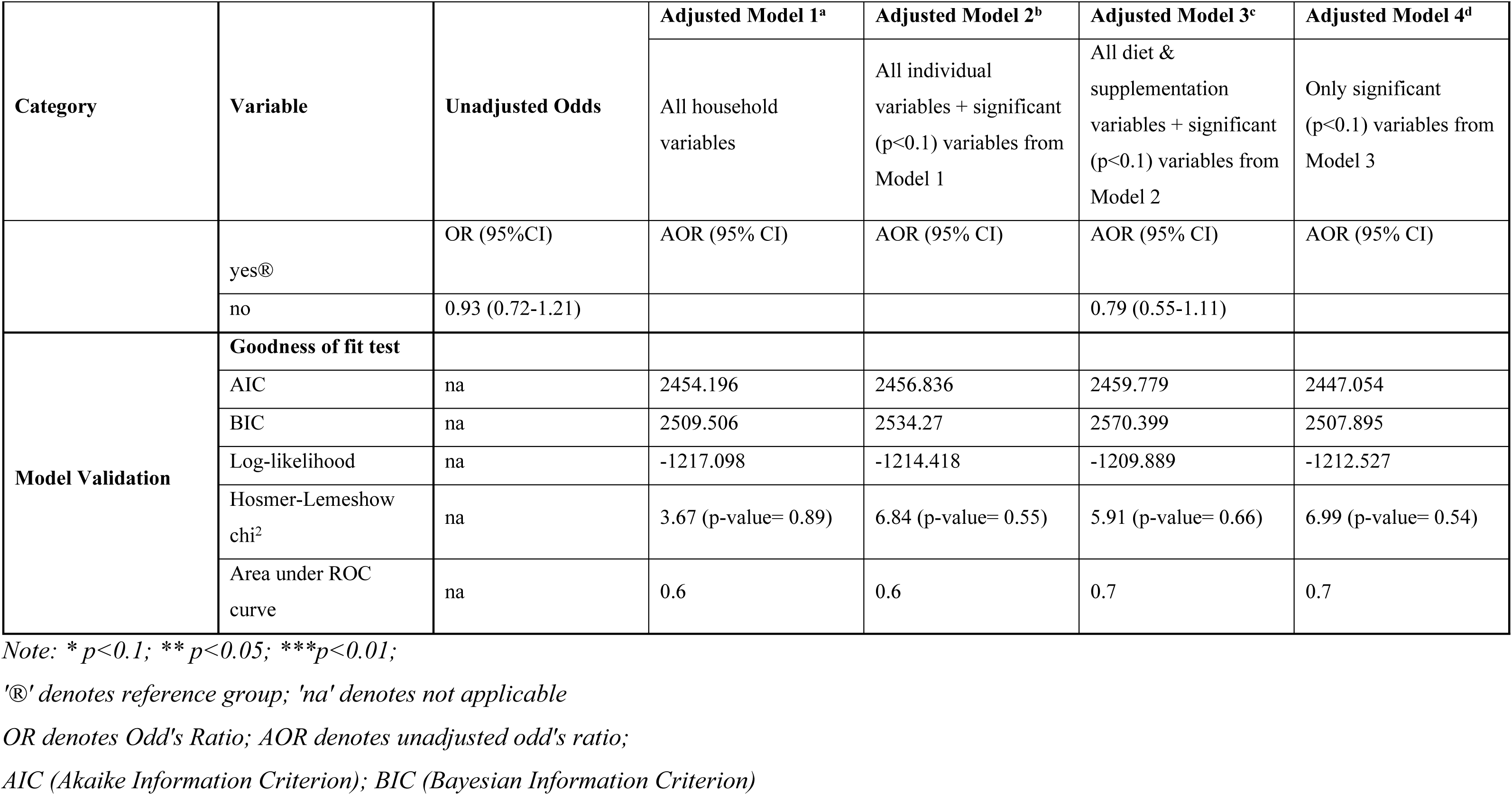
Findings from binary logistic regression analyses and goodness of fit tests.

### Sensitivity analysis

Fig 3 shows the predicator variables and the odds of anaemia from the adjusted regression model (model 4) from the main analysis and sensitivity analysis. The comparison reveals that same predictor variables were part of the model 4 in both the analysis, with minor deviation in the magnitude of odd’s ratio. In the model 4 of sensitivity analysis, consumption of pulses and beans does not show significant association with the odds of anaemia, while the age category, cooking method and consumption of fortified rice shows a pronounced significant association with odds of anaemia outcome. Detailed findings for sensitivity analysis are presented in Supplementary Table 1.

**Fig 3:**
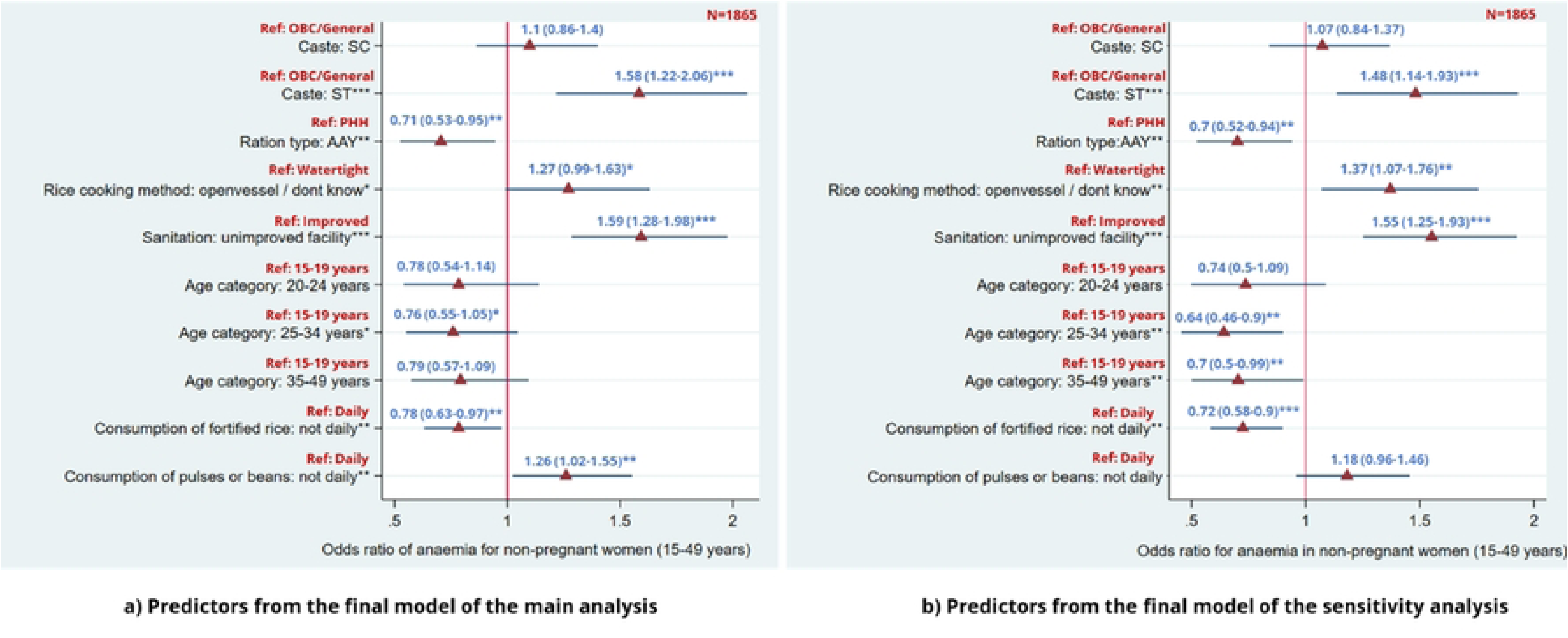
Comparison of Odds ratio from final model of main analysis and sensitivity analysis (Note: * p<0.1; ** p<0.05; ***p<0.01)

### Classification Analysis & Regression trees (CART)

For the CART analysis, the predictors from model 4 of the adjusted regression model were used as ‘circumstances’, while ‘prevalence of anaemia’ was considered as ‘outcome variable’. The formula considered for CART algorithm:

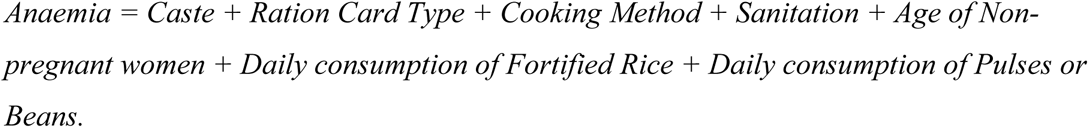

Fig 4 presents the tree obtained based on the CART analysis. The average prevalence of anaemia in the pooled data is 57%. The group of respondents (women 15-49 years) who most often experience anaemia **(furthest behind group)** have the following circumstance(s): Caste (ST) and Sanitation (not improved). The group’s prevalence is on average 72%. The group of respondents (women 15-49 years) who least often experience anaemia **(furthest ahead group)** have the following circumstance(s): Caste (SC/OBC/GEN), Sanitation (improved), Fortified Rice Consumption (daily) and Cooking Method (Watertight). The mean anaemia prevalence for this group is 39%.

**Fig 4:**
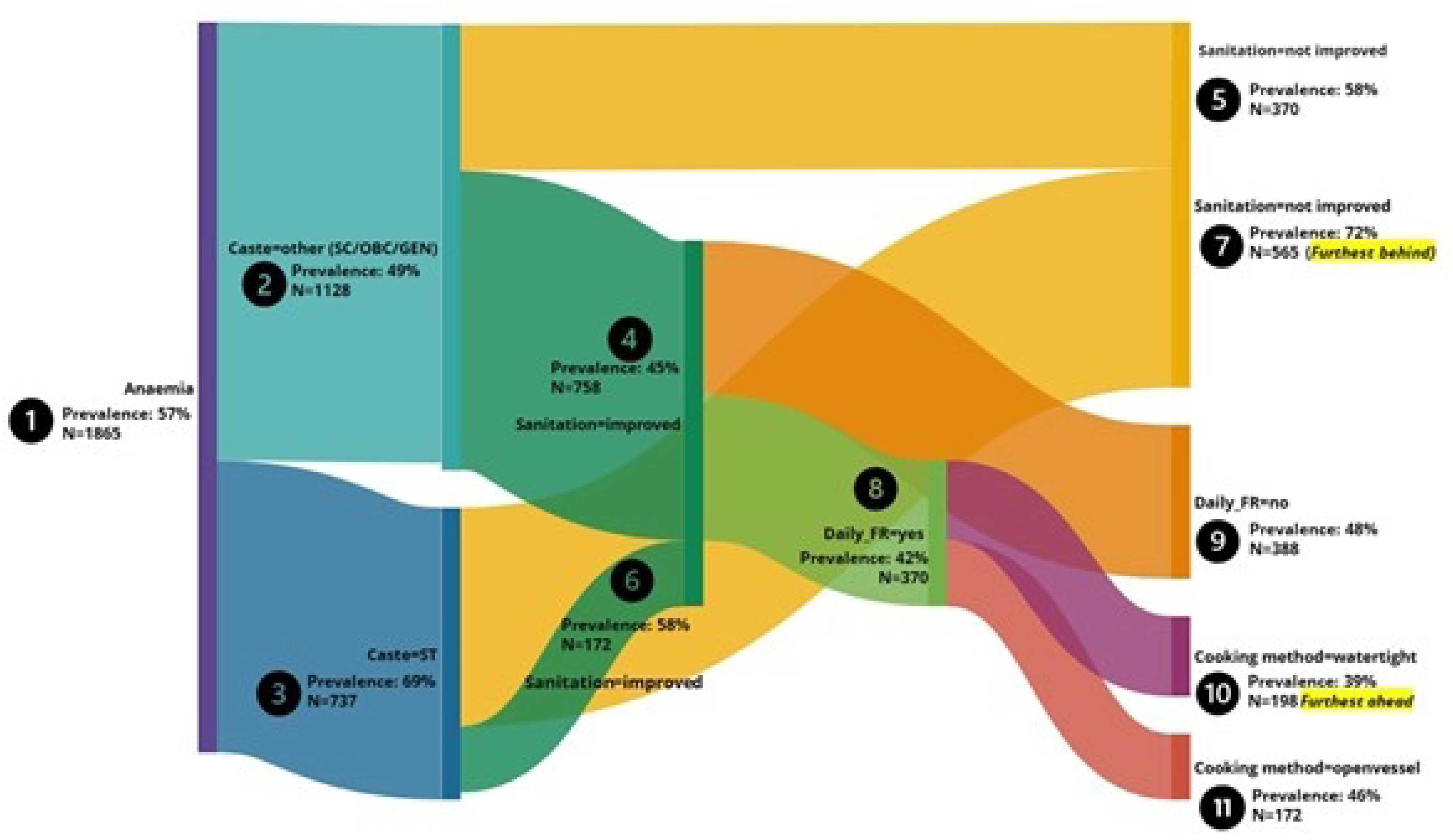
CART Tree for prevalence of anaemia

## Discussion

The paper attempted to fulfil four main objectives. First, it provided an overview of the rice fortification programme across two aspirational districts – Chandauli and Malkangiri in Uttar Pradesh and Odisha respectively, in terms of its scope, coverage and implementation post the roll out until the endline assessment, demonstrating a successful roll-out and scale-up of the rice fortification programme in both the districts despite varying local context and differences in supply chain management. Further, the household characteristics indicates stark socio-economic and demographic disparities between the two districts, with ST predominantly residing in Malkangiri and OBC and SC forming the majority in Chandauli. The significant gap in access to basic amenities like sanitation and clean fuel between the two districts reveals broader infrastructural challenges that contribute to health inequities, including anaemia. Unimproved sanitation is known to lead to parasitic infections, which damage intestinal walls, cause blood loss, and reduce the body’s ability to absorb nutrients from food (14). Malkangiri, with only 26.2% access to improved sanitation, lags far behind Chandauli, highlighting the urgent need to address sanitation as part of any anaemia reduction strategy. The disparity in access to clean fuel, with less than one-fifth of households in Malkangiri using it, further emphasizes the compounding effects of socio-economic deprivation on health outcomes. Page et al. (2015) in their study on pregnant women found that exposure to biomass smoke was linked to an increased risk of mild and moderate-to-severe anaemia, independent of other covariates (15). In a recent study conducted on non-pregnant women of reproductive age in Ethiopia, similar findings have been revealed (16). Exposure to unclean fuel triggers systemic inflammation through inflammatory cytokines that is known to cause anaemia (17).

Second, the comparison of anaemia prevalence between NFHS-5 and endline revealed that the roll out of rice fortification programme appears to have contributed to a decline in anaemia prevalence among non-pregnant women in both districts. The findings on reduction in prevalence of anaemia post consumption of iron-fortified food are consistent with many global and national studies (18–22). However, the reduction in anaemia prevalence was more pronounced in Chandauli (7.6 percentage points) compared to Malkangiri (4.6 percentage points). Despite the larger quantity of fortified rice being received by beneficiaries in Malkangiri, the decline is lower than that of Chandauli. Factors such as access to sanitation, drinking water, and healthcare, might be limiting the effectiveness of fortified rice intervention in Malkangiri. Moreover, the negligible change in median haemoglobin levels indicates that while fortification may help address moderate and severe anaemia, sustained intervention for longer period is required to observe an improvement in overall iron levels across the population. The surge in mild anaemia prevalence among non-pregnant women in both Malkangiri and Chandauli in the endline is possibly due to upward shift of moderate and severe anaemia cases to mild category.

Third, the findings from both the chi-square significance test and adjusted regression analysis underscore several critical factors influencing anaemia prevalence among non-pregnant women of reproductive age (15-49 years). The significant differences in anaemia prevalence between Malkangiri and Chandauli, with the former showing a much higher rate (69.7% versus 43.7%), highlight the geographical disparities in health outcomes that may be driven by variations in socio-economic conditions, access to health services, and dietary patterns. The higher prevalence in Malkangiri, a tribal-dominated region, reflects the vulnerability of these populations, as further evidenced by the elevated rates of anaemia among ST (68.9%), compared to SC (51.9%) and OBC/general categories (47%). The role of caste and other socio-demographic factors in determining anaemia prevalence is further supported by the adjusted regression analysis. Women from scheduled tribes had 58% higher odds of anaemia than women from OBC/general categories, indicating the persistent disadvantage faced by tribal communities in terms of nutritional outcomes. Access to basic services such as improved sanitation, clean drinking water, and clean cooking fuel emerged as strong determinants of anaemia. The higher anaemia prevalence among women from households without improved sanitation (66.5%) compared to those with improved facilities (47.4%) and the significant adjusted odds ratio of 1.59 suggest that poor sanitation exacerbates health vulnerabilities, likely through mechanisms related to frequent infections and poor hygiene. Similarly, the use of unimproved drinking water sources and unclean cooking fuel were associated with higher anaemia prevalence, underscoring the critical role of basic infrastructure in public health outcomes. Education was another key factor, with anaemia prevalence decreasing as educational attainment increased. Women with no schooling had the highest prevalence (63.8%), while those with secondary education (51.3%) and graduates (41.6%) fared better. The positive effect of education likely operates through multiple channels, including better health awareness, improved dietary choices, and greater access to healthcare services. These findings align with those of Sharif et al. (2023) (23), who analysed NFHS data from 2005 to 2021 and identified several socio-demographic factors such as low economic and educational status, rural residence, and higher childbearing rates as predictors of anaemia levels among different social groups of women in India. In addition, women from scheduled caste and scheduled tribes were more susceptible to anaemia compared to OBC and general category women, with the prevalence rate showing a slight increase from 2005–06 to 2019–21.

The current study also revealed interesting dietary patterns related to anaemia. Women who consumed pulses, beans, milk, curd, or fruits daily had lower anaemia prevalence, highlighting the importance of a balanced and diverse diet in combating micronutrient deficiencies. The findings are consistent with Jin et al (2022) (24), suggesting that a better-quality and diverse diet is linked to lower odds of anaemia among non-pregnant women of reproductive age. Our study also revealed that in populations affected by poor socio-economic conditions, inadequate living environments, and insufficient dietary intake, daily consumption of fortified rice may not fully translate into improved anaemia status.

Women consuming fortified rice daily were found to have a higher prevalence of anaemia compared to non-consumers (61.1% vs. 47.8%), despite its nutritional potential. Further analysis (Supplementary Table 2) revealed that these women were predominantly from highly vulnerable households—nearly half belonged to scheduled tribes, about 59% lacked access to improved sanitation, and 74% relied on unclean cooking fuel. Additionally, a significant proportion of these women had low educational levels, with about 50% having never attended school, and overall quality of diet was poor, with low consumption of nutrient-rich food groups. These findings suggest that the beneficial effects of fortified rice may have been negated by the cumulative burden of poverty, poor living conditions and poor dietary intake, rather than reflecting a physiological limitation of the fortified rice itself.

Moreover, the common practice of cooking fortified rice in open vessels with draining of excess water (reported by 82.7% of daily consumers) could potentially reduce the nutritional benefits, as it may lead to the loss of added micronutrients during cooking. Azam et al. (2021) (25), in their evaluation of various cooking methods on the loss of iron and zinc micronutrients in fortified and non-fortified rice, found that cooking rice in a rice cooker without washing it beforehand retained the highest levels of both iron and zinc in both rice types. This suggests that using the appropriate cooking method, particularly the watertight method, can help preserve more micronutrients, which can contribute to combating anaemia.

Interestingly, dietary habits commonly associated with better nutritional outcomes, such as regular consumption of fish, eggs, or meat, as well as frequent intake of millet, were found to be linked to higher anaemia prevalence. Further investigation reveals that this correlation may be attributed to the fact that most consumers belonged to Malkangiri district who face greater socio-economic disadvantages (in terms of poor access to improved sanitation, clean fuel and education among others) compared to those in Chandauli district. Despite the nutritional advantages of these food groups, the residents of Malkangiri remain more susceptible to anaemia to a greater degree, particularly due to persistence of basic structural inequalities (26).

Fourth, the CART analysis identified the most vulnerable groups, with scheduled tribes and women from households without improved sanitation emerging as the most at-risk populations, experiencing anaemia prevalence as high as 72%. This finding suggests that anaemia interventions must prioritize these groups and adopt a multi-sectoral approach that addresses not only food-based solutions like fortified rice but also broader socio-economic determinants of health, such as sanitation and access to clean water. Conversely, the group with the lowest prevalence of anaemia included women from other castes (SC/OBC/GEN), households with improved sanitation, daily consumers of fortified rice, and those using watertight cooking methods, with anaemia prevalence at just 39%. This reinforces the idea that a combination of improved living conditions and dietary practices is critical in reducing anaemia prevalence. Similar observations on integrated interventions have been made by researchers in the past to reduce the prevalence of anaemia (27–29).

Our study has several strengths. First, it contributes to the limited evidence on the effect of rice fortification interventions across districts in India by using primary data. Second, the endline assessment employed a sampling approach consistent with NFHS-5, including the method for haemoglobin testing, providing comparable statistics on various demographic, household, and individual factors influencing anaemia, specifically for a representative sample of non-pregnant women aged 15-49 years. Third, beyond significance testing and regression analysis, our study explores pathways using CART to identify the groups most and least affected by anaemia, an approach not previously undertaken by researchers.

One of the main limitations of this research is its cross-sectional survey design, which limits the ability to make causal inferences and only allows for the identification of associations between dependent and independent variables. Additionally, the baseline estimate (NFHS-5) covers both TPDS and non-TPDS beneficiaries, whereas the endline includes only TPDS beneficiaries. Furthermore, the endline study does not assess the quantity of food consumed by each target group, as it follows the NFHS-5 design, which only records food consumption frequency. This limitation affects the depth of the analysis and our understanding of dietary impacts on the target groups.

## Conclusion

The findings of this study have important implications for public health policy. First, while the rice fortification programme has demonstrated some success, particularly in reducing moderate and severe anaemia, it should be complemented with strategies to address sanitation, access to clean fuel, and dietary diversity. Interventions must be tailored to meet the specific needs of different socio-economic groups, with a focus on improving access to basic services for marginalized communities, particularly in tribal regions like Malkangiri.

Additionally, the findings suggest the need for enhanced monitoring and evaluation of the rice fortification process to ensure consistency and quality, as well as community-based education campaigns to increase awareness of dietary factors that enhance iron absorption. Finally, the government should consider expanding the fortification programme to include other staple foods, and integrating anaemia interventions with other social protection measures, such as health and nutrition services for women of reproductive age.

In conclusion, while the rice fortification programme has made a positive impact on reducing anaemia prevalence, particularly among the most vulnerable groups, addressing the structural inequities and socio-economic factors influencing anaemia remains critical and must be complemented by broader efforts to improve living conditions, access to basic services, and education to achieve sustainable improvements in women’s health.

## Data Availability

N/A

## Acknowledgement

We extend our gratitude to the state governments of Odisha and Uttar Pradesh for their cooperation in facilitating the assessment. We are also grateful to the team of TRIOs Development Support (P) Ltd. for their assistance in data collection and haemoglobin measurements, as well as to the respondents who provided consent and participated in the study. The authors also acknowledge and value the constructive feedback provided by the reviewers.

## Supporting Information

S1 Table: Logistic Regression Results for Sensitivity Analysis

S2 Table: Characteristics of women based on consumption of fortified rice

S3 Table 3: Spearman correlation matrix

